# Failed detection of the full-length genome of SARS-CoV-2 by ultra-deep sequencing from the recovered and discharged patients retested viral PCR positive

**DOI:** 10.1101/2020.03.27.20043299

**Authors:** Fengyu Hu, Fengjuan Chen, Yaping Wang, Teng Xu, Xiaoping Tang, Feng Li

**Author notes:** Correspondence: Xiaoping Tang, Feng Li, Guangzhou Eighth People’s Hospital, Guangzhou Medical University, 627 Dongfeng Dong Rd, Guangzhou 510000, China. These authors contributed equally (Fengyu Hu, Fengjuan Chen, Yaping Wang).

## Abstract

Over 10 percent of recovered and discharged patients retested positive for SARS-CoV-2, raising a public health concern whether they could be potential origins of infection. In this study, we found that detectable viral genome in discharged patients might only mean the presence of viral fragments, and could hardly form an infection origin for its extremely low concentration.

---

The novel coronavirus (SARS-CoV-2) infection seems to sweep across the globe (over 100,000 cases by March 7, 2020) ever since its outbreak. Fortunately, the majority of individuals infected with SARS-CoV-2 will recover with supportive therapy despite the absence of direct antiviral agents. However, some discharged patients retested to be viral RNA positive in Wuhan^1^. In Guangdong, 14 % of recovered and discharged patients retested positive for SARS-CoV-2 ^2^, thus raising a public health concern whether they could be potential origins of infection.

## Methods

This study followed the guideline of the Ethics Committee of Guangzhou Eighth People’s Hospital, which is appointed by the local government to treat patients suspected and confirmed to be infected by the SARS-CoV-2. Throat and anal samples were collected for viral RNA extraction and detection using reverse transcriptional real-time polymerase chain reaction (RT-PCR) as previously reported ^3^. In brief, two PCR primer and probe sets, which target orf1ab (FAM reporter) and N (VIC reporter) genes separately, were used to detect the presence of the SARS-CoV-2 sequence. Samples considered to be viral positive when either or both set(s) gave a reliable signal(s).

Metagenomic sequencing used a standard protocol as reported ^4^. Briefly, the libraries were generated after cDNA synthesis and subjected to sequencing on Illumina Nextseq using a single-end mode (1×75bp). Qualified viral sequence reads were aligned to the SARS-CoV-2 database. We conducted three levels of sequencing including standard (20 M reads), deep (60M reads), and ultra-deep (300M reads).

## Result

According to the regulation of Guangdong, 120 recovered individuals retested for the SARS-CoV-2 virus within 2 weeks after discharge by Feb 24, 2020, and 15 of them (12.5%) were RNA positive confirmed by local CDC and admitted to our hospital for quarantine. Throat and anal samples were collected for repeated detection. Among them, 10 patients had paired throat and anal samples for viral detection (Table. 1). Interestingly, we found that more anal samples (5 of 10) were positive than throat samples (2 of 10) in the same patients. However, the viral titers, if any, seemed very low as the Ct numbers were from 37 to 40, at the lowest detectable limit (cut off Ct=40-42), raising the suspicion of the absence of the complete SARS-CoV-2 virus.

**Table 1.** PCR and NGS sequencing result.

| Patient ID | Sample ID | Sample type | PCR result | SARS-CoV-2 Viral genome Reads /total |  |  |
| --- | --- | --- | --- | --- | --- | --- |
|  |  |  | Ct (N+Orf1ab) | 20 M reads | 60 M reads | 300 M reads |
| GZ8H-001 | 01 | Throat | Neg/39 | Neg | Neg | 5 |
|  | 02 | Anal | 38/Neg | 3 | 5 | 16 |
| GZ8H-002 | 03 | Throat | Neg/Neg | Neg | Neg | Neg |
|  | 04 | Anal | 36/37 | 2 | 6 | 28 |
| GZ8H-003 | 05 | Throat | Neg/Neg | Neg | Neg | Neg |
|  | 06 | Anal | 39/Neg | 4 | 4 | 9 |
| GZ8H-004 | 07 | Throat | Neg/Neg | Neg | Neg | Neg |
|  | 08 | Anal | Neg/Neg | Neg | Neg | Neg |
| GZ8H-005 | 09 | Throat | Neg/Neg | Neg | Neg | Neg |
|  | 10 | Anal | Neg/Neg | Neg | Neg | Neg |
| GZ8H-006 | 11 | Throat | Neg/Neg | Neg | Neg | Neg |
|  | 12 | Anal | Neg/Neg | Neg | Neg | Neg |
| GZ8H-007 | 13 | Throat | 40/Neg | 6 | 3 | 13 |
|  | 14 | Anal | 38/40 | 4 | 5 | 21 |
| GZ8H-008 | 15 | Throat | Neg/Neg | Neg | Neg | Neg |
|  | 16 | Anal | 37/38 | 14 | 18 | 36 |
| GZ8H-009 | 17 | Throat | Neg/Neg | Neg | Neg | Neg |
|  | 18 | Anal | Neg/Neg | Neg | Neg | Neg |
| GZ8H-010 | 19 | Throat | Neg/Neg | Neg | Neg | Neg |
|  | 20 | Anal | Neg/Neg | Neg | Neg | Neg |

Next, we checked whether the full-length SARS-CoV-2 virus was present with ultra-deep metagenomic sequencing technology. Our result showed a perfect match between metagenomic sequencing and PCR tests based on the observation that viral reads were only detected in PCR positive samples (01, 02, 04, 06, 13, 14, and 16). All the reads were mapped on the viral genome individually (Fig. 1). Interestingly, we found that even the ultra-deep sequence (30 times over the standard model) could not obtain enough reads to cover the full-length genome. The viral reads were scattered across the viral genome and no fixed pattern was observed between individuals or between different samples from the same individual.

**Figure 1.**
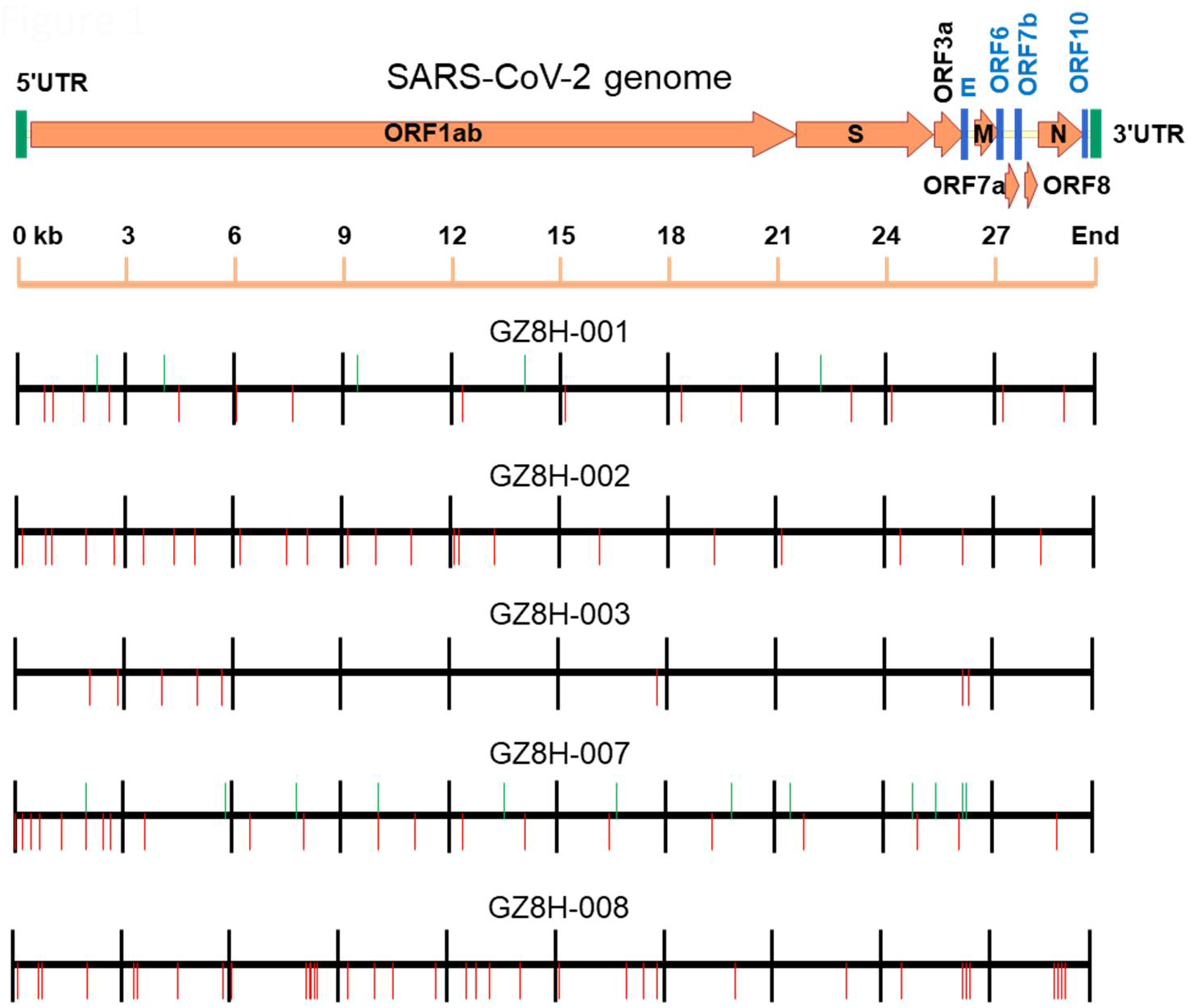
Reads distribution on SARS-CoV-2 genome. Line 1: SARS-CoV-2 genome organization, and the 5’ and 3’ untranslated terminal repeats (UTR, green) and open reading frame (ORF, dark yellow) are shown. Line 2: Scale of SARS-CoV-2 genome, kb: kilobase. Line 3-7: Distribution of viral reads obtained from ultra-deep metagenomic sequencing for patients GZ8H-001, 002, 003, 007, and 008, respectively. For each patient, reads from the throat sample are labelled upwards (Green line) and reads from the anal sample are labelled downwards (Red line) on the genome location.

## Discussion

Our study showed that a low level of fragment genome could be detected in the recovered and discharged patients, but mainly in the anal samples. Not all laboratory detections showed viral RNA positive (data not shown). As the SARS-CoV-2 entry receptor, ACE2 is widely expressed in multiple organs ^5^, we postulated that in certain types of cells infected by the SARS-CoV-2 virus, virus replication is incompletely suppressed even after clinical recovery for unknown reasons, resulting in the disconnectedly shedding of trace amount of zombie virus post-discharge.

Therefore, our result indicated that the detectable viral genome in discharged patients only means the presence of viral fragments and that an infection origin could hardly form for its extremely low concentration.

## Data Availability

The data used in this study is not available.

## Author contribution

Hu, Chen and Wang contributed equally to this study. Tang and Li contributed equally as senior authors.

Concept and design: Hu, Tang, Li.

Acquisition, analysis, or interpretation of data: Hu, Chen, Wang, Li.

Sequence and reads mapping: Xu.

Drafting of the manuscript: Hu, Li.

Supervision: Tang

## References

1. Lan L, Xu D, Ye G, et al. Positive RT-PCR Test Results in Patients Recovered From COVID-19. JAMA. 2020.

2. F K. 14% of recovered & discharged patients in Guangdong retested positive for Covid-19. 2020; https://www.theonlinecitizen.com/2020/02/26/14-of-recovered-discharged-patients-in-guangdong-retested-positive-for-covid-19/.

3. Chen W, Lan Y, Yuan X, et al. Detectable 2019-nCoV viral RNA in blood is a strong indicator for the further clinical severity. Emerg Microbes Infect. 2020;9(1):469–473.

4. Jing-Wen Ai, Hao-Cheng Zhang, Teng Xu, Wu J. Optimizing diagnostic strategy for novel coronavirus pneumonia, a multi-center study in Eastern China. medRxiv. 2020.

5. Hamming I, Timens W, Bulthuis ML, Lely AT, Navis G, van Goor H. Tissue distribution of ACE2 protein, the functional receptor for SARS coronavirus. A first step in understanding SARS pathogenesis. J Pathol. 2004;203(2):631–637.

